# Predicting longitudinal brain atrophy in Parkinson’s disease using a Susceptible-Infected-Removed agent-based model

**DOI:** 10.1101/2022.05.01.22274521

**Authors:** Alaa Abdelgawad, Shady Rahayel, Ying-Qiu Zheng, Christina Tremblay, Andrew Vo, Bratislav Misic, Alain Dagher

## Abstract

Parkinson’s disease (PD) is a progressive neurodegenerative disorder characterized by accumulation of abnormal isoforms of alpha-synuclein. Alpha-synuclein is proposed to act as a prion in PD: in its misfolded pathologic state it favours the misfolding of normal alpha-synuclein molecules, spreads trans-neuronally, and causes neuronal or synaptic damage as it accumulates. This theory remains controversial. We have previously developed a Susceptible-Infected-Removed (SIR) computational model that simulates the templating, propagation and toxicity of alpha-synuclein molecules in the brain. Here we test this model with longitudinal MRI collected over four years from the Parkinson Progression Markers Initiative (1068 T1 MRI scans, 790 PD, 278 matched controls). We find that brain deformation progresses in subcortical and cortical regions. The SIR model, using structural connectivity from diffusion MRI, recapitulates the spatiotemporal distribution of brain atrophy observed in PD. We show that connectome topology and geometry significantly contribute to model fit. We also show that the spatial expression of two genes implicated in alpha-synuclein synthesis and clearance, *SNCA* and *GBA*, also influences the atrophy pattern. We conclude that the progression of atrophy in PD is consistent with the prion-like hypothesis and that the SIR model is a promising tool to investigate multifactorial neurodegenerative diseases over time.

## INTRODUCTION

Parkinson’s disease (PD) is characterized by the pathological intracellular aggregation of misfolded alpha-synuclein (aSyn) into Lewy bodies and neurites (Dickson et al., 2009; Spillantini et al., 1997). In the brain, these deposits often appear in a stereotypical fashion, emerging in the olfactory bulb and caudal brainstem and then ascending towards the midbrain, limbic areas, and cerebral cortex (Braak et al., 2003, 2004). This spatiotemporal distribution patterns of pathology has led to the hypothesis that misfolded aSyn may harbor prion-like properties (Brundin & Melki, 2017), allowing it to spread between cells and impose its misfolded conformation onto native endogenous, otherwise normal aSyn from the recipient cell (Peng et al., 2020). Indeed, the injection of synthetic aSyn preformed fibrils or brain lysates from patients with a synucleinopathy has demonstrated the local formation of aSyn pathology and its propagation through brain networks in wild-type and transgenic mice, rats, and non-human primates (Henrich et al., 2020; Luk, Kehm, Zhang, et al., 2012; Masuda-Suzukake et al., 2013; Rey et al., 2016, 2018; Uemura et al., 2020; Watts et al., 2013).

In humans, the evidence for a prion-like behavior of pathological aSyn has so far been indirect. For instance, in patients who received fetal mesencephalic neuronal transplants, Lewy-related pathology could be observed inside cells that were grafted a decade earlier (Kordower et al., 2008; Li et al., 2008), suggesting that pathology spread to the grafts from the surrounding milieu. Also, using MRI-derived volume deformation and cortical thinning as proxy measures of tissue atrophy, the pattern of brain changes observed in *de novo* PD patients was shown to significantly overlap with the brain’s connectivity pattern (Pandya et al., 2019; Yau et al., 2018; Zeighami et al., 2015). However, other studies have shown that the distribution of aSyn pathology is not solely explainable by brain connectivity and that other cell-autonomous factors play a role in shaping aSyn pathology (Gonzalez-Rodriguez et al., 2020; Henrich et al., 2020; Surmeier et al., 2017). Thus, the mechanisms underlying the accumulation and propagation of pathological aSyn in PD remain unclear.

One way to understand these mechanisms is through computational modeling. We have recently developed an agent-based Susceptible-Infected-Removed (SIR) model that simulates the fate of individual aSyn proteins in the brain to recreate, based on cell-autonomous factors and brain connectivity, the atrophy pattern seen in PD (Zheng et al., 2019). The local factors in this case are expression of the genes *SNCA* and *GBA*, which we take as proxies of synthesis and clearance of aSyn. Using this model, we previously recreated the pattern of atrophy observed at baseline in *de novo* PD patients from the Parkinson’s Progression Markers Initiative (PPMI) cohort and demonstrated that both structural connectivity and gene expression were central to shaping the propagation of aSyn in the brain (Zheng et al., 2019). We subsequently used the same model to explain the propagation of pathologic aSyn injected into different brain regions of wild-type mice (Rahayel et al., 2021). Here we apply the model to longitudinal MRI data from PPMI.

We measured the progression of atrophy in PD patients over one, two, and four years and applied the agent-based SIR Model to assess if *SNCA* and *GBA* gene expression and structural features of the connectome significantly contributed to recreating the atrophy patterns. We found that the agent-based SIR Model accurately recreated the atrophy observed longitudinally in PD and that both gene and connectivity are significant contributors of atrophy.

## METHODS

### Participants

Longitudinal data from 709 PD patients and 279 healthy control participants were included from the PPMI database (www.ppmi-info.org), for a total of 1068 MRI scans and associated clinical measures. The PPMI is a longitudinal observational international study aimed at assessing progression markers of PD and includes a comprehensive set of clinical and MRI measures acquired in patients with de novo PD and healthy controls (Marek et al., 2011, 2018).

To be included in the PPMI, PD patients: 1) had at least two features among resting tremor, bradykinesia, and rigidity or either asymmetric resting tremor or asymmetric bradykinesia, 2) had a diagnosis of PD for less than two years, 3) had a baseline Hoehn and Yahr stage of I or II, 4) had a dopamine transporter binding deficit confirmed using SPECT scan, 5) were not expected to require medications for PD within six months of the baseline assessment, 6) were at least 30 years old, and 7) did not have dementia. For healthy controls, a Montreal Cognitive Assessment (MoCA) score below 27 or a first-degree relative with a clinical diagnosis of idiopathic PD led to exclusion. The longitudinal follow-up of PPMI now extends to approximately 5 years; for this study, only the participants with MRI acquisition performed at baseline and at either one, two, and/or four years were considered for analysis due to the limited number of scans acquired at three (3 participants) and five years (2 participants).

### Clinical Measures

At each visit, patients underwent the Movement Disorders Society-Unified Parkinson’s Disease Rating Scale (MDS-UPDRS) (Goetz et al., 2007) and a cognitive assessment that included the MoCA (Nasreddine et al., 2005), the Symbol-Digit Modalities Test, the Letter-Number Sequencing test, the Benton Judgment of Line Orientation test, the semantic and phonemic fluency tasks, and the total recall, delayed recall, and recognition tasks from the Hopkins Verbal Learning Test-Revised (Weintraub et al., 2015). Other clinical measures included the RBD Screening Questionnaire (Stiasny-Kolster et al., 2007), where a score ≥5 indicates probable REM sleep behavior disorder, the Geriatric Depression Scale (GDS), the State-Trait Anxiety Inventory (STAI), and the Scales for Outcomes in Parkinson’s Disease-Autonomic (SCOPA-AUT).

### MRI

#### MRI acquisition

T1-weighted MRI brain images were acquired at different sites across the United States, Canada, and Europe, with the following parameters: repetition time (TR) = 2,300 ms; echo time (TE) = 2.98 ms; field of view (FOV) = 256 mm; flip angle = 9; and voxel size = 1 mm^3^. The acquisition protocols are available on the PPMI website (http://www.ppmi-info.org/study-design/research-documents-and-sops/).

#### Deformation-based morphometry

Deformation-based morphometry (DBM) was performed on the baseline and longitudinal T1-weighted scans of PD patients and controls to derive whole-brain individual maps representing the deformation needed for a voxel to be normalized to the template space (using the MNI152-2009c template). DBM was done using the default parameters available in the CAT12 toolbox in SPM12 (www.neuro.uni-jena.de/cat). This resulted in a set of processed image files for each participant that included a voxel-wise whole-brain map of Jacobian determinants, which was used as the measure of local brain tissue atrophy after the application of a 2 mm full width at half maximum isotropic smoothing kernel. Images were visually inspected at each step and excluded if abnormal or if the automated quality rating was below 80%.

#### Brain parcellation

The normalized smoothed Jacobian determinants maps were next parcellated using a previously used atlas made of 42 cortical and subcortical brain regions from the left hemisphere for which regional *SNCA* and *GBA* expression as well as structural connectome features were available (Zheng et al., 2019). This atlas included 34 cortical regions derived from the Desikan-Killiany atlas (Desikan et al., 2006) and 7 subcortical regions, namely the putamen, caudate, pallidum, thalamus, hippocampus, amygdala, and accumbens, available as part of the FreeSurfer processing stream (http://surfer.nmr.mgh.harvard.edu). Due to its importance in PD, the substantia nigra was additionally included based on the segmentation available from a 7T MRI basal ganglia atlas (https://www.nitrc.org/projects/atag) (Keuken et al., 2014). Using FLIRT (https://fsl.fmrib.ox.ac.uk/fsl/fslwiki/FLIRT), the 42-region atlas was then linearly registered to the individual deformation maps and a set of 42 regional deformation values were extracted for each map using the MarsBaR region of interest toolbox for SPM (https://marsbar.sourceforge.net). Note that the atlas only included regions from the left hemisphere due to the *SNCA* and *GBA* gene expression for the right hemisphere being available for only 2 of the 6 post-mortem brains included in the Allen Human Brain Atlas (AHBA) (Hawrylycz et al., 2012). Moreover, we only applied the model to a single (left) hemisphere due to possible errors associated with the detection of interhemispheric white-matter connections using deterministic streamline tractography (see below).

#### Regional atrophy standardization

A W-score approach was then used to account for the normal effects of age and sex on brain morphometry (La Joie et al., 2012; Tremblay et al., 2021). The regional deformation values from each PD patient’s image were converted into age- and sex-corrected W-scores based on the values observed in the 157 controls available at baseline. There was no significant difference in age and sex between the controls (age: 60.1± 11.9; 66% male) at baseline and the PD group (BL age: 60.9± 10; 63% male – Y1 age: 60.9 ± 9.3; 63% male – Y2 age: 60.9 ± 9.3; 63% male – Y4 age: 64.4 ± 9.9; 69% male) at each time point. Only the values from controls seen at baseline were used for standardization due to the limited number of controls who underwent follow-up MRI. The standardization formula was:

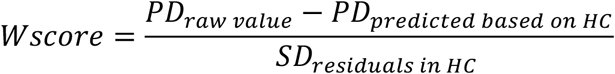

where the predicted value for a PD patient based on control data was given by (β_1_*age + β_2_*sex + β_3_). In other words, this yielded regional deformation values that represent the difference between a PD patient’s deformation value and the deformation value that is expected for their age and sex. W-scores are essentially Z-scores corrected for age and sex (La Joie et al., 2012). The individual W-scores were averaged between patients for a given region, resulting in a set of 42 regional W-scores for baseline and each of the three follow-up time points. The average W-score seen at each follow-up time point was then subtracted from baseline to yield a W-score difference over time (i.e., atrophy progression over one, two, and four years). A negative W-score difference represents atrophy progression in PD, whereas a positive W-score indicates volume expansion. The three sets of 42 atrophy difference values, one for the difference between baseline and every follow-up time point, were the observed patterns of atrophy progression to which we compared the pattern of simulated atrophy generated *in silico* by the agent-based SIR Model.

### Agent-based SIR Model

#### Overview of the model

The agent-based SIR Model simulates the brain spread of aSyn based on regional *SNCA* and *GBA* gene expression and inter-regional connectivity (Zheng et al., 2019). In this model, the synthesis and degradation of aSyn agents are modulated by the local expression of *SNCA* and *GBA*, respectively. Every agent in a brain region can belong to one of three compartments: “Susceptible” when representing the normal protein, “Infected” when representing the misfolded protein, and “Removed” when the protein gets degraded or spreads to another region. Note that in this model all infected agents are also deemed to be infectious. Every Susceptible agent can turn into an Infected agent when it encounters an Infected agent in a region. Both Susceptible and Infected agents have a probability of either being degraded inside a region or to spread to a connected area. The probability of spreading to another region is based on the strength of the connectivity (i.e., streamline density, see below) between the source and the target regions.

The model is run by first initiating pathology inside a seed region, here the substantia nigra, and simulating the spread over a total of 10,000 iterations. At each iteration, a simulated atrophy value is generated for every region. Atrophy is assumed to result from the combined effects of local accumulation of infected agents and deafferentation. To investigate how well the parameters of the spreading model replicated the progression of atrophy in PD, the spread was simulated with the same 42-region atlas used for the MRI-derived observed patterns of atrophy. This allowed us to compare the pattern of regional values of simulated atrophy to the patterns of atrophy progression observed between baseline and one, two, and four years. In other words, following the initiation of pathology at the substantia nigra, the model used information about structural connectivity and regional gene expression to modulate the behavior of aSyn agents in order to simulate local accumulation of aSyn pathology and atrophy. The model was implemented as five different modules, namely the production of normal aSyn, the clearance of normal and misfolded aSyn, the misfolding of normal aSyn, the propagation of normal and misfolded aSyn, and the accrual of atrophy (see below for details about each module).

#### Production of normal aSyn

In the model, the synthesis of aSyn inside every region was modulated based on the regional gene expression of *SNCA*, which was extracted using the software toolbox abagen (Markello et al., 2021), available at https://abagen.readthedocs.io/, for the 42 regions based on the six post-mortem brains of the AHBA (Hawrylycz et al., 2012). The values were averaged across samples to yield an expression vector of synthesis that was inserted back into the model (Zheng et al., 2019). The synthesis rate in region i is given by the probability of new agent synthesis per unit time, αi:

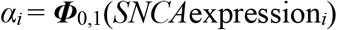

where Φ0,1(·) is the normal cumulative distribution function. The increment of normal agents in region i is given by αiSiΔt, where Δt is the total time and Si is the region size. The time increment used for the main analyses was set at Δt = 0.1, but peak correlation fits were robust with values from 0.1 to 0.9 (see Supplementary Figure 1).

#### Clearance of normal and misfolded aSyn

Likewise, the degradation of aSyn inside every region was modulated based on the regional gene expression of *GBA*, which was also extracted from the AHBA. The clearance rate of both normal and misfolded agents in region *i* per unit time occurred with the probability distrbution *β*_*i*_:

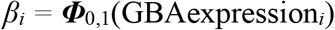

where ***Φ***_0,1_(·) is the normal cumulative distribution function. The probability of an agent still being active after total time *Δt* is given by *lim*_*δτ*→0_(1 − *βδτ*)^Δ*t*/*δτ*^ = *e*^−*β*Δ*τ*^. In other words, as the degradation rate increases, the probability of an agent to remain active in the region decreases. Accordingly, the proportion of cleared agents within timestep *Δt* is 1 – *e*^−*β*Δ*t*^.

#### Misfolding of normal aSyn (infection transmission)

Infected agents have the ability to promote misfolding of susceptible agents and turn them into infected agents. The probability of a susceptible agent that survived clearance of not being infected is 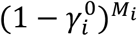, where *M*_*i*_ is the population of infected agents in region *i* and 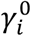 is the baseline likelihood that a single misfolded agent turns a susceptible agent into an infected agent. The baseline likelihood 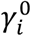 is given by 1/*S*_*i*_, where *S*_*i*_ is the region size. Accordingly, the probability per unit of time that a susceptible agent surviving clearance in region *i* turns into an infected agent due to the action of at least one of the *M*_*i*_ infected agents present in region *i* is given by 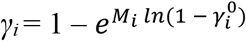. As for the previous module, the probability that a susceptible agent remains susceptible after total time *Δt* is given by 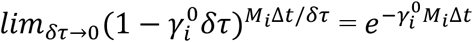, whereas the probability that a susceptible agent becomes infected after total time *Δt* is given by 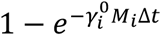. As a result, the increment of the population of normal proteins *N*_*i*_ in region *i* is:

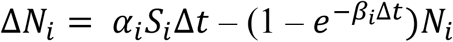

After each timestep, the populations of susceptible (Ni) and infected agents (Mi) are respectively updated as follows:

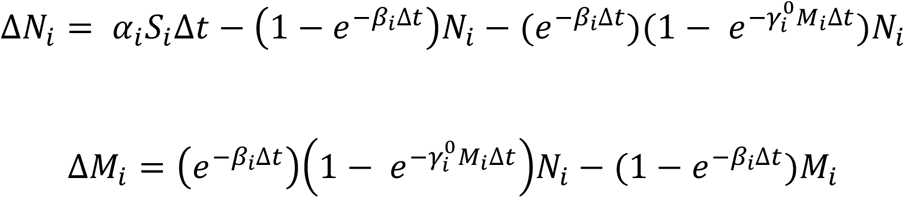

#### Propagation of normal and misfolded aSyn

Every susceptible and infected agent has a probability to spread to other brain regions. To implement this, the structural connectivity matrix created previously for the 42-region atlas was used (see Zheng et al., 2019). Briefly, this was created using 1,027 preprocessed diffusion-weighted and T1-weighted MRI images from the Human Connectome Project (2017 Q4; 1,200-subject release). The diffusion data were reconstructed in the individual T1-weighted image space using generalized q-sampling imaging. Voxel-wise quantitative anisotropy and the spin distribution function were measured to assess the density of water diffused in different directions. Deterministic streamline tractography was then performed for each region using DSI Studio (www.nitrc.org/projects/dsistudio), resulting in 100,000 streamlines per region with the following parameters: angular cut-off of 55, step size of 0.5 mm, minimum length of 20 mm, and maximum length of 400 mm. For the purpose of the agent-based model, the connectivity strength between each seed-target region pair was defined as the density of streamlines (i.e. streamline count) between the two regions normalized by the target region size and the mean length of streamlines.

To account for the mobility pattern of an agent between regions, we used a distance matrix and a structural connectivity matrix. The distance matrix was constructed by calculating the Euclidean distance of corresponding streamlines between every pair of regions. For the structural connectivity matrix, a connection profile based on the density of streamlines was created for each region with self-connection set to 0; then concatenated to form a 42×42 structural connectivity matrix for each subject. Finally, a group-consensus approach was adopted by averaging 35 % of the most commonly occurring edges across all subjects to generate one group-level structural connectivity matrix. To test for robustness, the analyses were also performed using different matrix densities (Supplementary Table 1).

Using the matrix of structural connectivity, every agent can either remain in region *i* or enter the edges (fiber tracts) with probabilities:

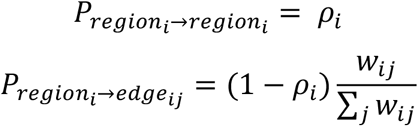

where *w*_*ij*_ is the undirected connection weight between region *i* and region *j* and *ρ*_*i*_ is the probability of an agent to remain in region *i*. This probability was set to 0.5 for every region. The choice of *ρ*_*i*_ led to negligible differences when simulating atrophy (Supplementary Figure 1). Likewise, both susceptible and infected agents can exist in an edge *(i,j)* or exit the edge per unit time with probabilities:

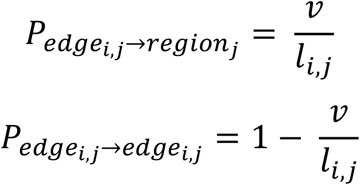

where *l*_*ij*_ is the length (Euclidean distance) of the edge between regions *i* and *j* and *v* is the propagation speed. The increments in quantity of normal and misfolded agents *N*_*i*_ and *M*_*i*_ in region *i* after a total time Δ*t* are as follows:

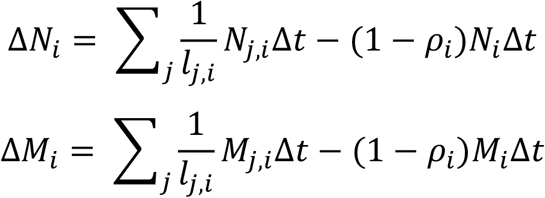

where *N*_*i,j*_ and *M*_*i,j*_ represent the populations of normal and infected agents in the edge between regions *i* and *j* respectively. *N*_*i,j*_ and *M*_*i,j*_ are updated as follows:

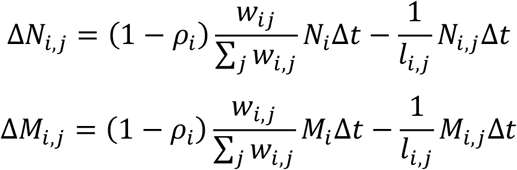

#### Accrual of atrophy

Tissue loss was modeled as the result of two processes: the direct toxicity from the accumulation of infected agents in region *i* and the deafferentation due to neuronal death in regions connected with region *i*. The incremental atrophy at time *t* over *Δt* in region *i* is given by:

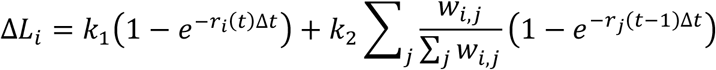

where *r*_*i*_(*t*) represents the proportion of misfolded agents in region *i* at time *t, k*_1_ is the weight (impact) of aSyn accumulation on tissue loss, and *k*_2_ is the weight (impact) of deafferentation from neighboring regions on tissue loss. Both *k*_*1*_ and *k*_*2*_ were set to 0.5 such that accumulation of infected agents and deafferentation had equal effect on the growth of the atrophy simulated by the model.

### Statistical analyses

#### Demographics and clinical variables

The demographics and clinical variables were compared between PD patients and controls at baseline and between PD patients at each follow-up time point versus baseline. Student’s two-sample t-tests and Mann-Whitney U tests were respectively used for normally and non-normally distributed continuous variables. Chi-square tests were used for comparing groups on categorical variables.

#### Longitudinal progression of atrophy

To examine the progression of brain atrophy in PD patients, we performed linear mixed-effect modeling to investigate if the effect of time was significant over the regional deformation values at each time point, namely at baseline and after one, two, and four years of follow-up. This resulted in a set of 42 separate models, one for each brain region. The random intercept was assigned at the patient level, while the fixed effect was the interaction of time with the age-and-sex corrected w-score DBM maps. The Benjamini-Hochberg procedure was used to control the false discovery rate (Benjamini et al., 2001) and a regional deformation change was considered significant when the *p* value was below 0.05.

#### Fit between observed and modeled pathology

The SIR model was run for a total of 10,000 iterations after injecting pathology into the substantia nigra. The propagation speed, *v*, which models the protein spreading rate, was set to 1. To check for robustness, variation in propagation speed values ranging from 0.1 to 10 resulted in negligible difference on the model fit (Supplementary Figure 1). Model fit between simulated and observed atrophy was measured using Spearman’s rank coefficient correlations. First, we investigated if the atrophy simulated in every region was significantly associated with the deformation value observed at baseline. At every time point (i.e., after one, two, and four years of follow-up), the regional simulated data was correlated with the regional observed atrophy difference, which is calculated by subtracting baseline W-score DBM value from that of follow-up time point. The peak fit between simulated and atrophy difference patterns observed between baseline and each time point corresponded to the highest correlation coefficient between the two metrics. To confirm that significance testing was not affected by spatial autocorrelation of brain atrophy, we used the toolbox BrainSMASH (Burt et al., 2020) to generate 1000 surrogate brain maps preserving spatial autocorrelation (for each comparison), as null distributions for assessment of statistical significance (Markello & Misic, 2021).

#### Null models

To investigate the impact of gene expression and connectivity on the spread of pathologic aSyn, we generated null models in which gene expression or connectivity were randomized. We then computed peak fits between the observed and simulated atrophy for each null model, and compared the spatial patterns thus obtained to the true peak fits between true and simulated atrophy. For the connectome null models, the impact of topology and/or geometry was investigated using both rewired and repositioned null models. In rewired null networks, using the Maslov-Sneppen algorithm in the Brain Connectivity Toolbox (sites.google.com/site/bctnet), pairs of connectivity strength between brain regions were randomly shuffled in the structural connectivity matrix while preserving the network’s original degree sequence and density; the rewiring per edge parameter was set to 100. In repositioned null networks, the spatial position of regions was randomly shuffled while preserving the network’s original degree sequence and connection profile. For gene expression null models, the values of either *SNCA* or *GBA* regional expression were randomly reassigned to each region. In every case, the shuffled connectivity or gene expression data were inserted back into the model and used to simulate the spread of agents. For each of the four types of null models (i.e., rewired, repositioned, *SNCA*, and *GBA* null models), the randomization was repeated 500 times to generate distributions of null peak fits. The original peak fit between the observed and simulated atrophy patterns was then compared using one-sample t-tests to the average peak fit distributions of the null models.

## RESULTS

### Participants

A total of 1,068 T1-weighted scans: 790 PD and 278 healthy controls were obtained from the PPMI cohort. Of these, 199 scans were rejected: 193 failed quality control, and 6 scans were acquired outside the follow-up time points investigated in this study (i.e., 3 and 5 years after baseline). This yielded a total of 869 scans from 238 HC and 631 PD. The PD scan numbers are 318 at baseline, 120 at one year, 108 at two years and 85 at four years. Only patients with a scan acquired at baseline and at least one follow-up time point were kept for further analysis, leaving samples of 113 patients between baseline and one year, 104 patients between baseline and two years, and 79 patients between baseline and four years. Only the complete sample of 157 healthy controls at baseline were included here due to the small number of follow-up scans.

There were no significant age, sex, and education differences at baseline between patients and controls (Table 1). PD patients had significantly higher scores on the MDS-UPDRS-I, MDS-UPDRS-II, MDS-UPDRS-III, GDS, and SCOPA-AUT, a higher percentage of probable RBD, and lower scores on the MoCA, Symbol-Digit Modalities Test, and the total recall, delayed recall, and recognition tasks from the Hopkins Verbal Learning Test-Revised. In PD patients, scores gradually worsened at each follow-up time point on the MDS-UPDRS-I, MDS-UPDRS-II, MDS-UPDRS-III, and the SCOPA-AUT.

**Table 1.**
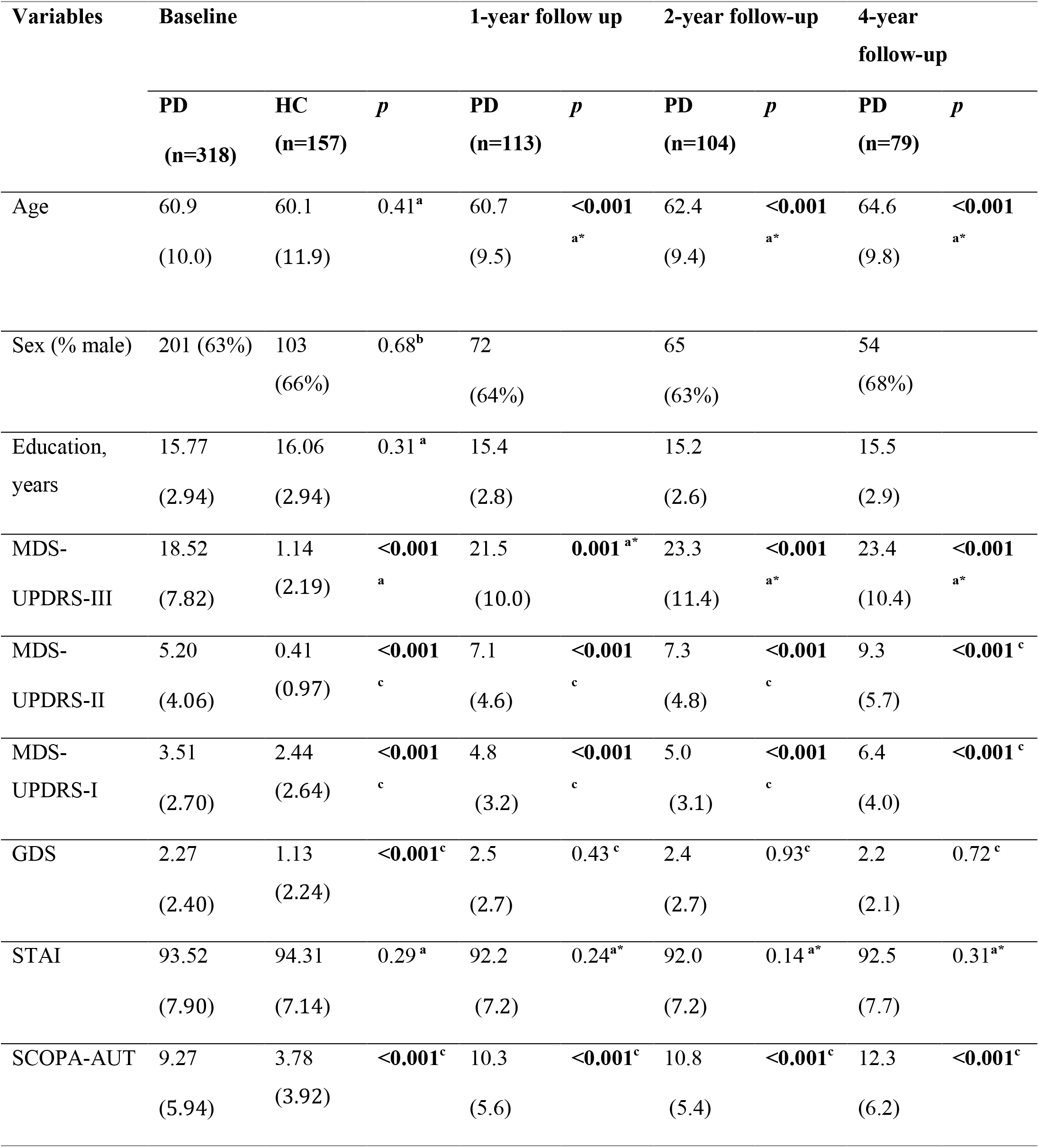
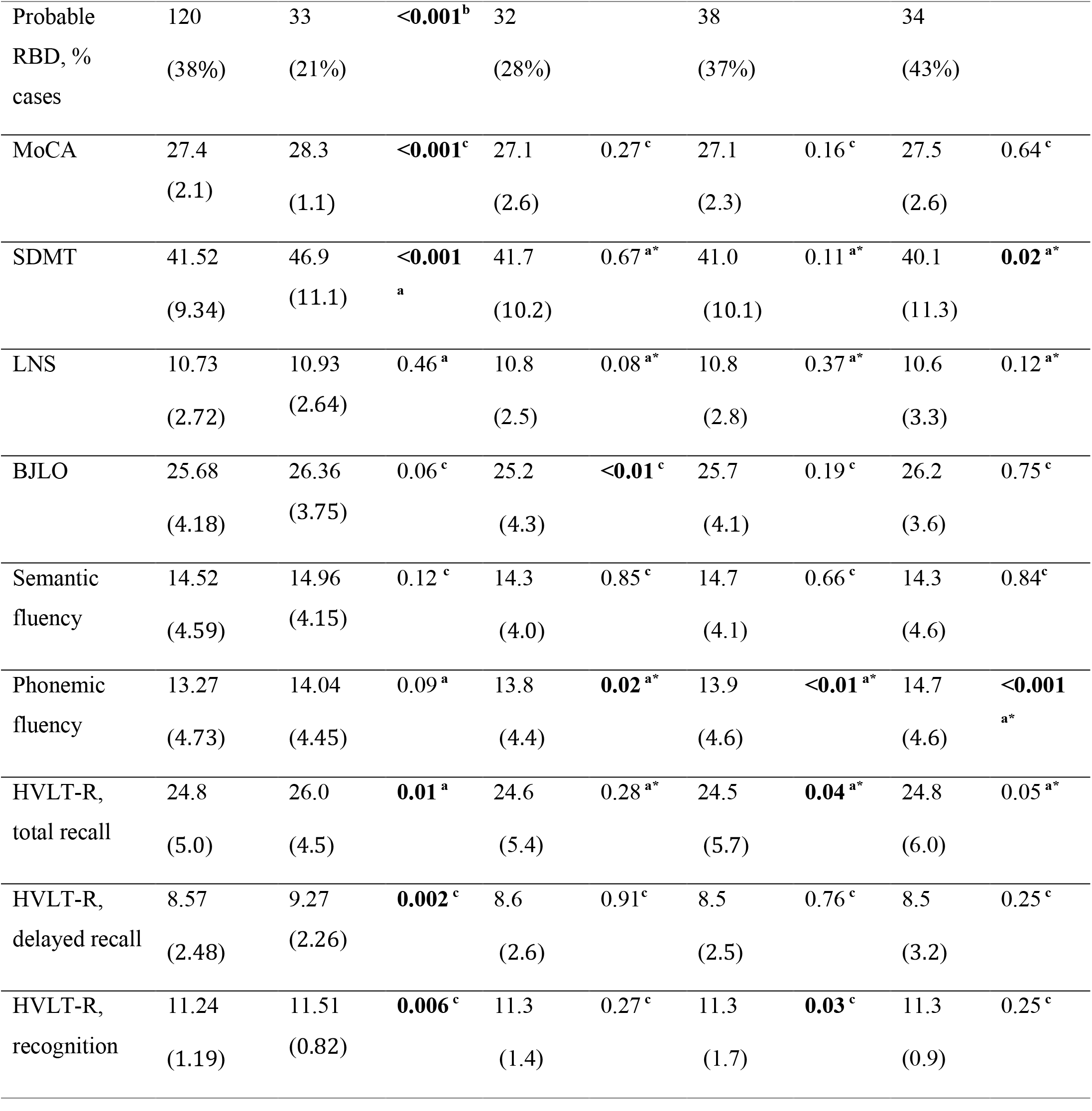
Demographics and clinical characteristics of patients and controls. Data are shown as mean (standard deviation). The performance in PD patients at the follow-up time point was statistically compared to their performance at baseline. ^**a**^ unpaired t-test, ^**a\***^paired t-test, ^**b**^chi-square test, ^c^Mann-Whitney U test. BJLO = Benton Judgment of Line Orientation; GDS = Geriatric Depression Scale; HC = healthy controls; HVLT-R = Hopkins Verbal Learning Test-Revised; LNS = Letter-Number Sequencing; MDS-UPDRS = Movement Disorders Society-Unified Parkinson’s Disease Rating Scale; MoCA = Montreal Cognitive Assessment; PD = Parkinson’s disease; RBD = REM sleep behavior disorder; SCOPA-AUT = Scales for Outcomes in Parkinson’s Disease-Autonomic; SDMT = Symbol-Digit Modalities Test; STAI = State-Trait Anxiety Inventory.

### Brain atrophy progresses over 4 years in PD

Using linear mixed-effect models, 23 of the 42 left hemisphere brain regions showed significant progression of deformation in PD over four years, while controlling for age and sex (Figure 1 and Supplementary Table 2). Specifically, between baseline and year one, atrophy increased in 14 regions, including the striatum, the temporal areas (i.e., middle and inferior temporal cortices, entorhinal cortex, parahippocampal gyrus, banks of the superior temporal sulcus, lingual and fusiform gyri), the isthmus of the cingulate cortex, the precuneus and inferior parietal cortex, the lateral occipital cortex, and the lateral orbitofrontal cortex. After two years of follow-up, the rostral anterior cingulate cortex, supramarginal cortex, temporal pole, and insula also start showing significant deformation. After four years of follow-up, atrophy was now additionally present in the posterior cingulate cortex, superior parietal cortex, superior temporal cortex, nucleus accumbens, and amygdala. Unlike the other regions, the insula demonstrated tissue expansion (Figure 1) both at baseline and increasing with time. This tissue expansion may represent an increase in CSF volume in the perisylvian area. These results confirm previous analyses of this dataset using slightly different methodology (Tremblay et al., 2021).

**Figure 1.**
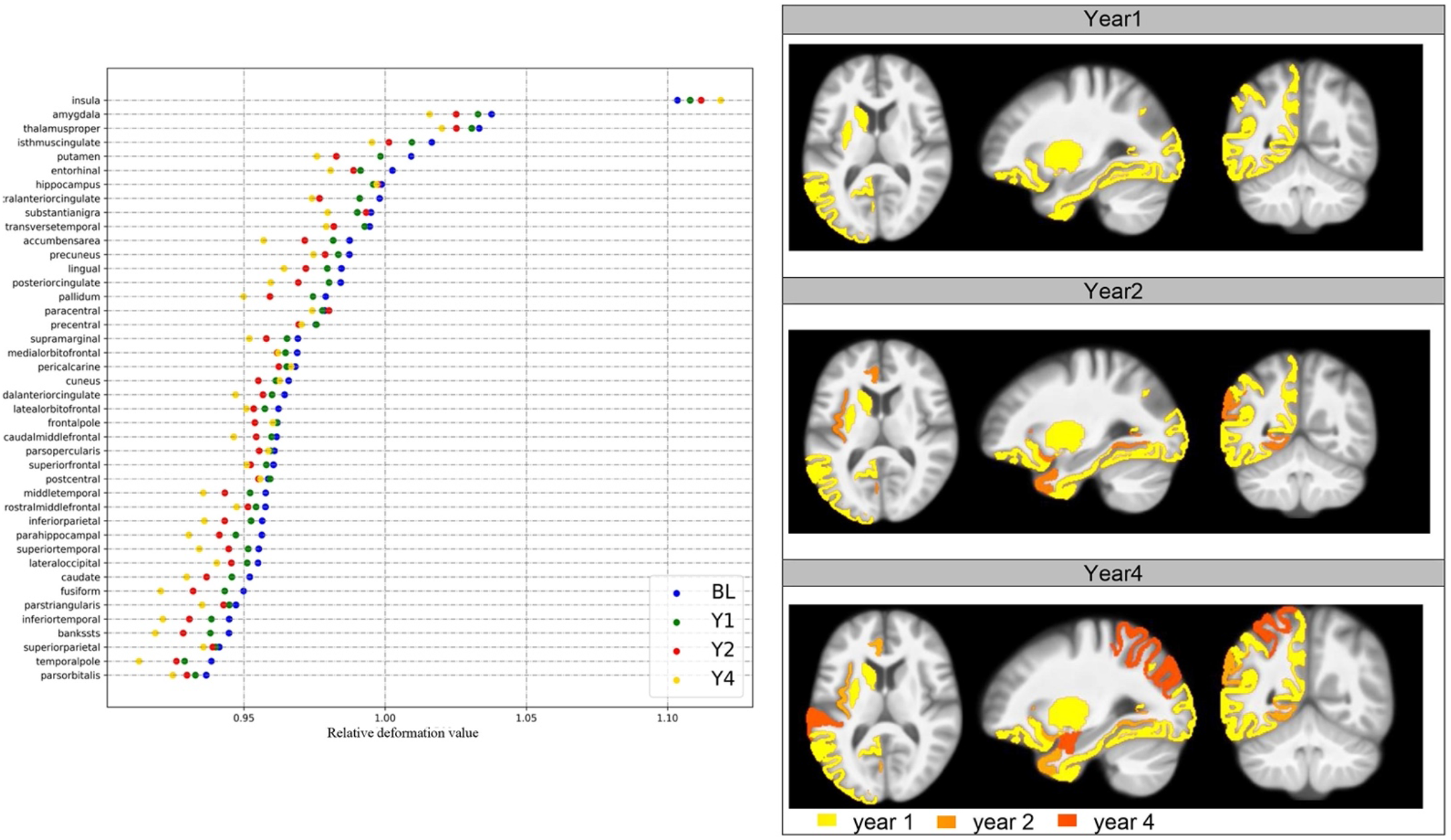
Regional longitudinal changes in PD over 4 years. **(A)** represents the DBM maps observed at baseline and during follow-up time points (i.e., one, two, and four years) in patients with PD for the 42 regions. **(B)** Brain maps showing the regions that were significantly deformed at each time point compared to baseline. Only the left hemisphere is shown due to limitations regarding the gene expression scores and the structural connectivity measures. Color bar reflects the first occurrence of a time effect on volume. Dark yellow, orange and red represent significant changes after 1, 2 years and 4 years of follow-up, respectively.

### The agent-based SIR Model recreates atrophy progression

Next, we used the agent-based SIR Model to simulate the spread of aSyn in the 42 regions and compared the pattern of atrophy simulated by the model to the patterns of atrophy progression observed in PD patients between baseline and one year, two years, and four years of follow-up. We found that the atrophy pattern simulated by the model significantly recreated the atrophy progression patterns observed longitudinally in PD (Figure 2). Specifically, the peak correlation between the simulated and observed patterns of atrophy at baseline was r=0.58 (p<0.0001) and occurred early during the spread of agents (i.e., timestep 500) (Figure 2A). The peak correlation between the simulated atrophy pattern and the progression of atrophy in PD was r=0.34 (p=0.03) at one year (Y1) and r=0.33 (p=0.03) at two years (Y2, Figure 2B); in contrast to the atrophy seen at baseline, the peak correlation fit was reached at much later timesteps (i.e., between timesteps 7000 and 9000), once the system has reached its equilibrium state. Similar results were obtained from significance testing using null models to account for spatial autocorrelation (Baseline: r = 0.62, p = 0.001, Y1: r = 0.33, p = 0.03, Y2: r = 0.29, p = 0.056). The simulated atrophy generated by the model did not recreate the pattern of atrophy progression seen between baseline and four years (r=0.2 p=0.1, Figure 2C).

**Figure 2.**
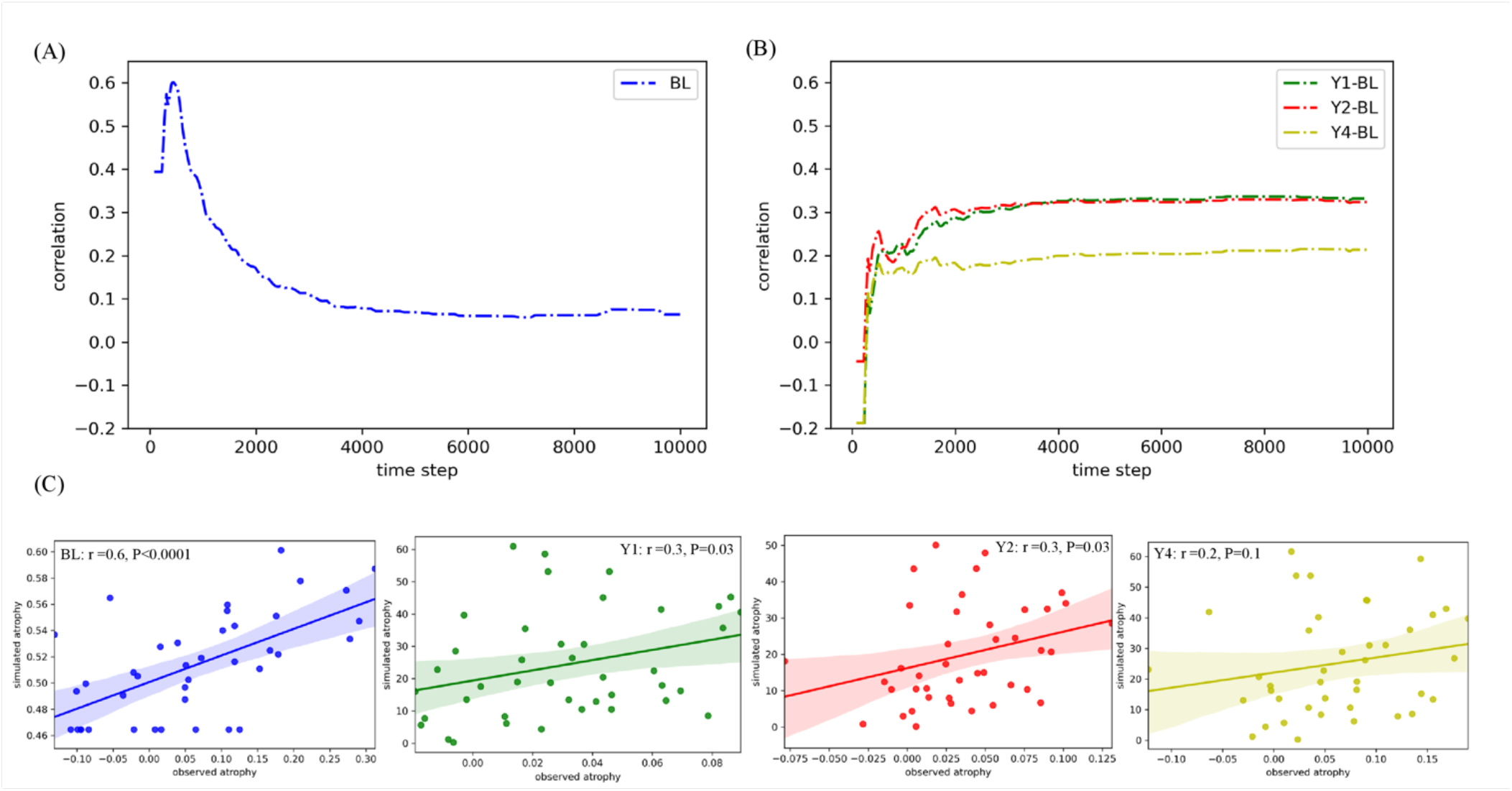
The agent-based SIR Model recreates the progression of brain atrophy. **(A)** The peak fit was assessed using Spearman’s rank correlation coefficient at each of the 10,000 simulation timestep between simulated pattern of atrophy to the patterns of atrophy observed at baseline and **(B)** atrophy difference at each follow-up time point (i.e., one, two, and four years). **(C)** Scatterplots showing the observed and simulated atrophy for each region at each simulation peak correlation fit.

To further confirm these results, we repeated the same analyses using structural connectivity matrices containing 30%, and 40% of the most occurring edges instead of the 35% density used for the main findings. Results were similar (Supplementary Table 1). Taken together, this demonstrates that the agent-based SIR Model recreates the progression of brain atrophy taking place over two years in PD.

### Brain connectivity shapes the progression of atrophy

To investigate if the connectome’s architecture shaped the progression of atrophy in PD, we generated sets of 500 rewired and 500 repositioned null models in which the connectome’s topology or geometry was randomized. For rewired models the peak fits were always significantly lower than the peak fit obtained with the true connectivity matrix (r_null_ ∼ 0.12, p<0.0001 at every time point; Figure 3A). Using repositioned models to randomize the physical position of brain regions, we also observed that the peak fit was significantly disrupted at every time point (r_null_ ∼0.29, p<0.0001 at every time point; Figure 3C). Taken together, this demonstrates that the brain’s connectivity pattern and spatial embedding shape the progression of brain atrophy in PD. The main findings here are reported for 35% connectome density; we found similar results when using different network densities (Supplementary Figure 1).

**Figure 3.**
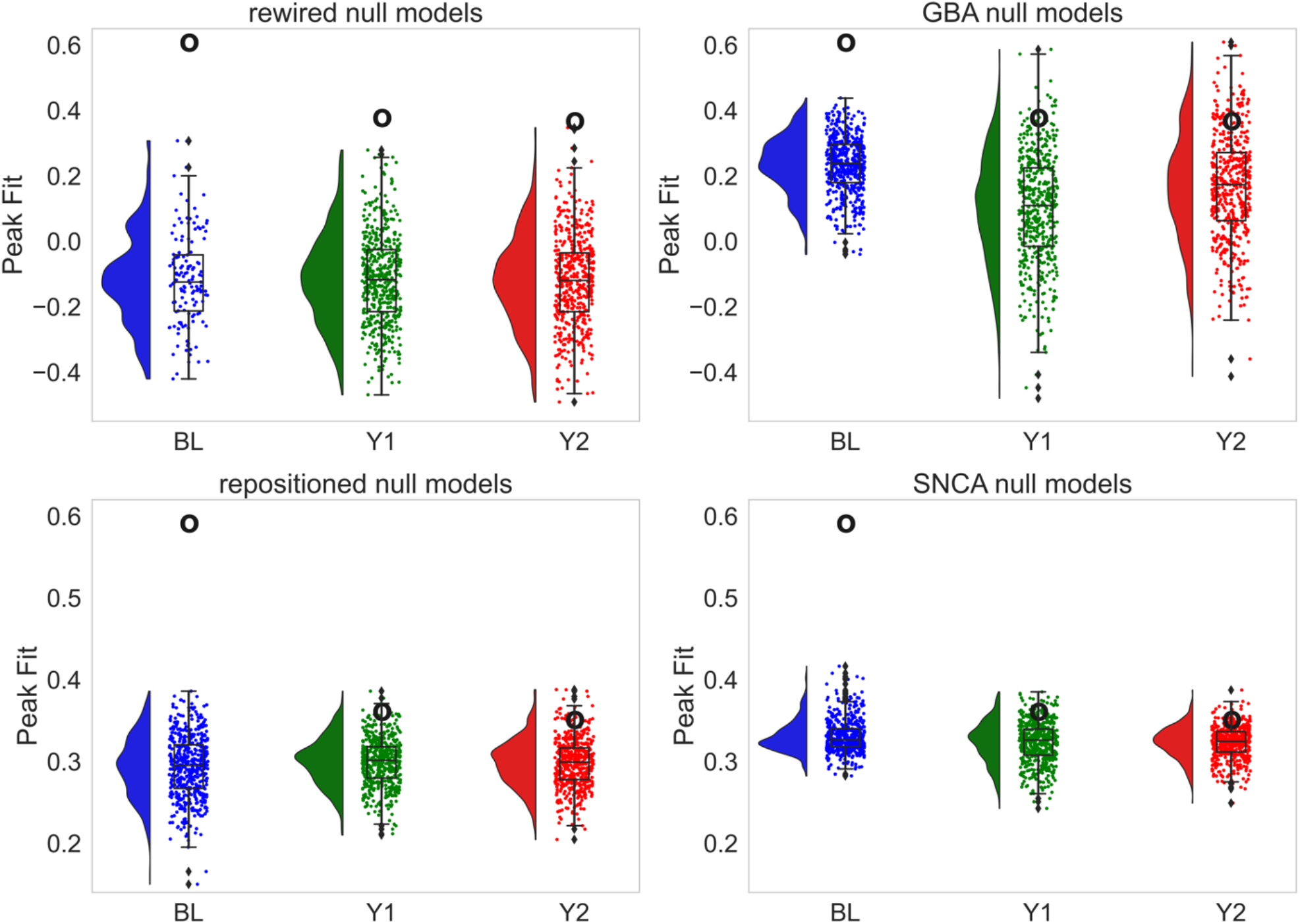
Brain connectivity and gene expression shape the progression of brain atrophy in PD. the distributions of null peak correlation fits when shuffling randomly either the **(A)** connectivity weights between regions, **(B)** the local expression of *GBA*, **(C)** the spatial embedding of regions, or **(D)** the local expression of *SNCA*. The average null peak fit was compared to the original peak fit obtained when using the real parameter. The black circle refers to the value of the peak correlation fit between the observed pattern of atrophy and the simulated pattern with the original non-shuffled parameter. The comparisons are made at baseline and for the one- and two-year time points. For all null models, there was a significant difference between the original fit and the shuffled fits at p<0.0001 using one-tailed t test.

### Gene expression shapes the progression of atrophy

To investigate if regional gene expression shaped the progression of atrophy in PD, the expression of *SNCA* or *GBA* was randomized across brain regions. The fit between the simulated and observed patterns of atrophy was significantly disrupted at baseline and at each of the following time points when randomizing *SNCA* (*SNCA*: r_null_=0.33 p<0.0001at baseline, r_null_=0.32 at one year, and r_null_=0.32 at two years with p<0.0001; Figure 3D) or *GBA* (r_null_=0.23 at baseline, r_null_=0.09 at one year, and r_null_=0.17 at two years with p<0.0001; Figure 3B).

## DISCUSSION

The prion-like model of PD makes three predictions: 1) misfolded aSyn isoforms act as a template to misfold normal aSyn molecules; 2) abnormal aSyn molecules propagate trans-synaptically via the connectome; 3) accumulation of misfolded aSyn leads to tissue damage in vulnerable regions. Here we test all three aspects of this model by simulating the fate of aSyn agents in the brain and comparing the resulting patterns to empirically derived atrophy maps from T1 MRI scans in people with PD.

We studied the progression of brain atrophy in newly-diagnosed PD patients over 4 years. Three main observations were made: first, as demonstrated previously with this dataset (Tremblay et al. 2021), atrophy increased significantly over four years, being found in the striatum early on and involving a greater number of cortical regions as disease progresses. Second, the SIR model recreated *in silico* the pattern of atrophy observed longitudinally in PD patients. Third, the SIR model demonstrated that both cell-autonomous factors like *SNCA* and *GBA* gene expression levels and non-cell autonomous factors, namely the topology and geometry of the connectome, shaped the spatiotemporal progression of atrophy. These findings further support the theory of PD as a propagating synucleinopathy.

Using age and sex-corrected measures of brain deformation, we found that 55% of brain regions showed significant atrophy in PD at some point over the 4-year follow-up. The regions with the strongest progression of atrophy over 4 years were the putamen and caudate and the middle and inferior temporal cortices. Atrophy in cortical regions such as the rostral anterior cingulate cortex and supramarginal cortex appeared after two years, whereas the limbic structures (i.e., amygdala and nucleus accumbens) and the superior parietal, posterior cingulate, and superior temporal cortices started showing atrophy after 4 years of follow-up. This atrophy pattern involving basal ganglia first, followed by mostly posterior cortical regions, was also described in a large meta-analysis from the ENIGMA consortium (Laansma et al., 2020), which included PD patients with overall more advanced disease than our *de novo* cohort. Note also that ENIGMA included the PPMI MRI scans used here although they only account for 15% of the 2357 PD datasets in that study. Interestingly, the substantia nigra, in which signal changes have been associated with the parkinsonian motor signs and symptoms associated with PD (Gaurav et al., 2021), was atrophied at baseline but did not show any atrophy progression during the follow-up years, suggesting that this region may have already reached a floor effect at the time of clinical diagnosis, at least in terms of volume deformation.

PD is pathologically characterized by the accumulation of misfolded aSyn in Lewy bodies and Lewy neurites (Spillantini et al., 1997). Two theories currently exist to explain the aSyn-related pathogenesis in the brain: the prion-like protein propagation and the regional vulnerability hypotheses (Brundin & Melki, 2017; Surmeier et al., 2017). According to the prion-like hypothesis, pathologic aSyn imposes its misfolded conformation onto native proteins that can then spread trans-synaptically between neurons, a hypothesis that is supported by several studies in animals (Luk, Kehm, Carroll, et al., 2012; Masuda-Suzukake et al., 2013; Rahayel et al., 2021). More recently, MRI studies performed in humans have shown that the pattern of atrophy observed in *de novo* patients with PD overlaps with known structural and functional networks (Pandya et al., 2019; Zeighami et al., 2015), suggesting that brain connectivity is a critical determinant of atrophy in synucleinopathies. However, there is also evidence that the connectivity alone does not completely explain the pattern of Lewy-related pathology, and that intrinsic factors may govern the selective vulnerability of certain regions or cell types (Fu et al., 2018; Gonzalez-Rodriguez et al., 2020). While several factors relating to cellular energetics and neurotransmitter metabolism have been proposed (Giguère et al., 2018), the concentration of normal aSyn and the expression of *SNCA* are also markers of cell vulnerability (Luna et al., 2018).

Our model explicitly incorporates normal aSyn production and breakdown, and randomizing these values degrades its ability to replicate observed atrophy, as does randomizing connectivity values. Applied to de novo PD patients, the SIR model has previously demonstrated that *SNCA* and *GBA* expression and brain connectivity both significantly influence the distribution of atrophy in the brain of PD patients (Zheng et al., 2019). Furthermore, the same model was recently used to predict the spread of pathologic aSyn injected in different brain regions of wild-type mice (Rahayel et al., 2021). However, no study had yet applied the agent-based model to the analysis of atrophy progression in PD.

More specifically, we found that the increase in brain atrophy observed at 1 and 2 years was significantly recreated by the model. The use of null networks in which either gene expression or brain connectivity were randomized shows that the atrophy depends upon both connectivity and aSyn synthesis and metabolism. This is in line with similar studies showing that pathology and atrophy occur along brain networks in other neurodegenerative diseases such as frontotemporal dementia (Brown et al., 2019; Shafiei et al., 2022; Zhou et al., 2012), Alzheimer’s disease (Raj et al., 2015; Vogel et al., 2020, 2021) and amyotrophic lateral sclerosis (Meier et al., 2020). In contrast to other computational models, which generally simulate the spread of abnormal proteins by relying on a connectivity-based diffusion mechanism, our agent-based model generates a pattern of propagation and atrophy that also takes local vulnerability into account by simulating the synthesis and degradation of aSyn as individual agents.

The progression of atrophy occurring after four years could not be replicated by the model. This may be due to the lower number of scans acquired at the four-year time point (85 versus 105 at two years) causing reduced statistical power. Also, attrition bias may be present whereby the group of PD patients still in the study at year 4 had milder disease (Tremblay et al., 2021). Another possibility is that neuron and synapse loss over time modified the patients’ connectivity structures, leading to inaccuracies in modeling spread of pathology using a healthy connectome. Future studies could integrate measures of ongoing loss of connectivity and integrate this into the SIR model.

Regional aSyn concentration was modulated in the SIR model to assess regional vulnerability to pathology accumulation. Shuffling the expression level of either *SNCA* or *GBA* resulted in significantly disrupted fit between observed and simulated data across all time points, suggesting the importance of expression of both genes in shaping the spatial pattern of disease spread longitudinally. In other words, regional variations in synthesis and clearance of aSyn, as indexed by *SNCA* and *GBA* expression, contribute to the PD atrophy progression pattern in our model. This is consistent with the fact that mutations in both genes are risk factors for genetic forms of PD (Gan-Or et al., 2018; Konno et al., 2016; Riboldi & Di Fonzo, 2019), and both are identified in genome wide associations studies of sporadic PD (Nalls et al., 2019).

Similarly, the randomization of connectome topology (via rewired null models) and spatial organization (via spatial null models) resulted in disrupted fit between observed and simulated data. While this is consistent with trans-neuronal propagation of a toxic agent, it can also be explained by other forms of connectivity-related co-atrophy. For example, interconnected areas may share local properties that render them similarly vulnerable to neurodegeneration. These include glucose metabolism, gene expression, neuronal cell count and shape, synaptic spine density, and other cytoarchitectonic features (Fulcher & Fornito, 2016; Richiardi et al., 2015; Scholtens et al., 2014), which may all influence local vulnerability.

This study has some limitations. First, the PD patients recruited as part of the PPMI study are younger and have less cognitive impairment than the more general population of PD patients (Marek et al., 2011). However, PPMI represents the largest longitudinal dataset of PD patients with MRI acquisition and clinical assessments. Second, the agent-based SIR model did not account for the cell loss that may have occurred as agents spread throughout the system; the atrophy occurring during the spread of agents may have modified the constraints of the system and therefore its outputs. For example, the substantia nigra is a source of propagating aSyn in our model having both high *SNCA* expression and widespread connectivity (Zheng et al., 2019); however, cell loss in this region could impact disease pattern in later stages of PD, something our model does not incorporate. Gene expression was only investigated for *SNCA* and *GBA* due to their known importance in aSyn synthesis and degradation; future studies should perform a more thorough evaluation of the different genes that may impact aSyn spread. Finally, our model does not consider the synergy between aSyn accumulation and autophagy-lysosomal dysfunction or mitochondrial failure (Hou et al., 2020; Senkevich & Gan-Or, 2020), which may also display regional variance.

In conclusion, we showed that brain atrophy progresses in PD over four years into patterns that could be recreated by the agent-based SIR model, a computational model that generates in silico the propagation of aSyn and brain atrophy using gene expression and connectivity. This computational model may represent a promising tool for better understanding the mechanisms underlying the progression of atrophy in neurodegenerative diseases.

## Data Availability

All code and data produced in the present study are available at https://github.com/alaaabdel/Longitudinal_DBM_Data and https://github.com/yingqiuz/SIR_simulator

https://github.com/yingqiuz/SIR_simulator

## ACKNOWLEDGMENTS

Alaa Abdelgawad reports Jeanne-Timmins Costello Fellowship – McGill University. Shady Rahayel reports a scholarship from the Fonds de recherche du Québec – Santé (FRQS). Alain Dagher acknowledges funding from the Canadian Institutes of Health Research (CIHR), the Michael J. Fox Foundation, the W. Garfield Foundation, the Alzheimer’s Association, and Healthy Brain for Healthy Lives Initiative (McGill University).

## DATA AVAILABILITY STATEMENT

W-score deformation maps at each year can be found at https://github.com/alaaabdel/Longitudinal_DBM_Data. The SIR model simulator can be accessed at https://github.com/yingqiuz/SIR_simulator. Any further data is available from the corresponding author upon request.

## AUTHORSHIP

All co-authors have been involved in drafting the article and revising it critically for intellectual content. All co-authors have read and approved the final version of the manuscript.

