## Supporting Information for "Predicting longitudinal brain atrophy in Parkinson’s disease using a Susceptible-Infected-Removed agent-based model"

| Time point | 40% density | | | 35% density | | | 30% density | | |
|---|---|---|---|---|---|---|---|---|---|
| | r | timestep | p-value | r | timestep | p-value | r | timestep | p-value |
| BL | 0.58 | 457 | $6.39e^{-5}$ | 0.58 | 500 | 1.24e-5 | 0.53 | 260 | $3.2e^{-4}$ |
| BL versus Y1 | 0.31 | 6417 | 0.04 | 0.34 | 8533 | 0.03 | 0.33 | 9354 | 0.03 |
| BL versus Y2 | 0.31 | 6417 | 0.04 | 0.33 | 7182 | 0.03 | 0.33 | 3252 | 0.03 |

**Supplementary Table 1. The model recreates atrophy at different network densities.** The peak Spearman's correlations between the simulated pattern of atrophy and the patterns of atrophy observed at baseline and at the one- and two-year time points remain when simulating the spread using network densities containing the top 30%, 35%, and 40% connections of the structural connectivity matrix. Timestep refers to the simulation timestep at which peak fit occurs. BL: baseline; Y1: year one; Y2: year two.

| | Year 1 | | | | | Year 2 | | | | | Year 4 | | | | |
|---|---|---|---|---|---|---|---|---|---|---|---|---|---|---|---|
| **Region** | **Coeff** | **SE** | **p** | **95% CI** | | **Coeff** | **SE** | **p** | **95% CI** | | **Coeff** | **SE** | **p** | **95% CI** | |
| Lateral orbitofrontal | -0.052 | 0.023 | **0.049** | -0.096 | -0.008 | -0.073 | 0.023 | **0.006** | -0.119 | -0.028 | -0.103 | 0.026 | **0.0004** | -0.154 | -0.053 |
| Pars orbitalis | -0.035 | 0.021 | 0.17 | -0.076 | 0.006 | -0.031 | 0.021 | 0.24 | -0.073 | 0.011 | -0.048 | 0.024 | 0.087 | -0.094 | -0.002 |
| Frontal pole | 0.014 | 0.034 | 0.76 | -0.052 | 0.081 | -0.051 | 0.035 | 0.24 | -0.12 | 0.018 | -0.039 | 0.039 | 0.43 | -0.115 | 0.037 |
| Medial orbitofrontal | -0.019 | 0.021 | 0.47 | -0.06 | 0.022 | -0.024 | 0.021 | 0.37 | -0.066 | 0.018 | -0.049 | 0.024 | 0.081 | -0.095 | -0.003 |
| Pars triangularis | 0.002 | 0.017 | 0.94 | -0.031 | 0.035 | 0.026 | 0.017 | 0.22 | -0.008 | 0.06 | 0.016 | 0.019 | 0.52 | -0.022 | 0.053 |
| Pars opercularis | 0.011 | 0.013 | 0.52 | -0.015 | 0.037 | 0.013 | 0.014 | 0.44 | -0.014 | 0.04 | 0.026 | 0.015 | 0.17 | -0.004 | 0.055 |
| Rostral middle frontal | -0.037 | 0.022 | 0.19 | -0.08 | 0.007 | -0.022 | 0.023 | 0.44 | -0.067 | 0.023 | -0.046 | 0.025 | 0.14 | -0.095 | 0.004 |
| Superior frontal | -0.008 | 0.018 | 0.76 | -0.043 | 0.027 | 0 | 0.018 | 0.99 | -0.036 | 0.036 | -0.028 | 0.02 | 0.26 | -0.068 | 0.011 |
| Caudal middle frontal | 0.009 | 0.014 | 0.63 | -0.018 | 0.036 | -0.002 | 0.014 | 0.94 | -0.029 | 0.026 | -0.009 | 0.016 | 0.69 | -0.039 | 0.022 |
| Precentral | 0.017 | 0.016 | 0.43 | -0.016 | 0.049 | -0.002 | 0.017 | 0.94 | -0.035 | 0.032 | 0.004 | 0.019 | 0.89 | -0.032 | 0.041 |
| Paracentral | 0.001 | 0.014 | 0.94 | -0.025 | 0.028 | -0.006 | 0.014 | 0.76 | -0.034 | 0.22 | -0.02 | 0.016 | 0.3 | -0.051 | 0.01 |
| Rostral anterior cingulate | -0.03 | 0.015 | 0.103 | -0.059 | 0 | -0.036 | 0.016 | **0.048** | -0.067 | -0.006 | -0.075 | 0.017 | **0.00011** | -0.109 | -0.042 |
| Caudal anterior cingulate | -0.012 | 0.01 | 0.33 | -0.032 | 0.008 | 0 | 0.01 | 0.98 | -0.02 | 0.021 | 0.006 | 0.12 | 0.703 | -0.017 | 0.029 |
| Posterior cingulate | -0.017 | 0.017 | 0.44 | -0.051 | 0.017 | -0.019 | 0.018 | 0.41 | -0.054 | 0.016 | -0.054 | 0.02 | **0.017** | -0.093 | -0.016 |
| Isthmus of cingulate | -0.057 | 0.016 | **0.001** | -0.088 | -0.025 | -0.051 | 0.017 | **0.0075** | -0.083 | -0.018 | -0.119 | 0.018 | **<0.0001** | -0.155 | -0.083 |
| Postcentral | 0.013 | 0.016 | 0.53 | -0.019 | 0.045 | -0.009 | 0.017 | 0.7 | -0.042 | 0.024 | -0.036 | 0.019 | 0.109 | -0.073 | 0.001 |
| Supramarginal | -0.035 | 0.017 | 0.08 | -0.067 | -0.002 | -0.044 | 0.017 | **0.029** | -0.077 | -0.01 | -0.077 | 0.019 | **0.00032** | -0.114 | -0.04 |
| Superior parietal | -0.2 | 0.016 | 0.33 | -0.052 | 0.012 | -0.027 | 0.017 | 0.19 | -0.061 | 0.006 | -0.72 | 0.019 | **0.0007** | -0.109 | -0.035 |
| Inferior parietal | -0.049 | 0.017 | **0.01** | -0.082 | -0.016 | -0.065 | 0.017 | **0.00087** | -0.098 | -0.031 | -0.142 | 0.019 | **<0.0001** | -0.18 | -0.105 |
| Precuneus | -0.047 | 0.014 | **0.002** | -0.074 | -0.021 | -0.047 | 0.014 | **0.003** | -0.075 | -0.02 | -0.134 | 0.015 | **<0.0001** | -0.164 | -0.103 |
| Cuneus | -0.022 | 0.013 | 0.163 | -0.083 | 0.151 | -0.013 | 0.013 | 0.44 | -0.046 | 0.013 | -0.022 | 0.014 | 0.202 | -0.051 | 0.006 |
| Pericalcarine | -0.012 | 0.017 | 0.59 | -0.047 | 0.022 | 0.007 | 0.018 | 0.79 | -0.029 | 0.042 | 0.022 | 0.02 | 0.39 | -0.017 | 0.061 |
| Lateral occipital | -0.044 | 0.014 | **0.007** | -0.071 | -0.016 | 0.015 | 0.008 | **0.021** | -0.067 | -0.01 | 0.016 | | **<0.0001** | -0.126 | -0.063 |
| Lingual | -0.036 | 0.015 | **0.042** | -0.07 | -0.006 | -0.03 | 0.016 | 0.11 | -0.061 | 0.001 | -0.085 | 0.017 | **<0.0001** | -0.119 | -0.051 |
| Fusiform | -0.06 | 0.022 | **0.018** | -0.104 | -0.017 | -0.083 | 0.023 | **0.0013** | -0.13 | -0.038 | -0.18 | 0.025 | **<0.0001** | -0.229 | -0.13 |
| Parahippocampal | -0.077 | 0.029 | **0.02** | -0.13 | -0.02 | -0.089 | 0.03 | **0.0084** | -0.15 | -0.031 | -0.16 | 0.33 | **<0.0001** | -0.22 | -0.095 |
| Entorhinal | -0.074 | 0.025 | **0.011** | -0.12 | -0.024 | -0.108 | 0.026 | **0.00032** | -0.16 | -0.056 | -0.12 | 0.029 | **<0.0001** | -0.18 | -0.063 |
| Temporal pole | -0.068 | 0.03 | 0.051 | -0.126 | -0.01 | -0.073 | 0.031 | **0.043** | -0.13 | -0.013 | -0.13 | 0.034 | **0.0007** | -0.2 | -0.064 |
| Inferior temporal | -0.074 | 0.02 | **0.001** | -0.113 | -0.036 | -0.065 | 0.02 | 0.0051 | -0.105 | -0.025 | -0.17 | 0.022 | **<0.0001** | -0.21 | -0.12 |
| Middle temporal | -0.065 | 0.017 | **0.001** | -0.099 | -0.031 | -0.081 | 0.018 | **<0.0001** | -0.12 | -0.046 | -0.18 | 0.02 | **<0.0001** | -0.22 | -0.14 |
| Superior temporal sulcus | -0.031 | 0.012 | **0.018** | -0.054 | -0.009 | -0.039 | 0.012 | **0.0037** | -0.063 | -0.016 | -0.041 | 0.013 | **0.0057** | -0.067 | -0.016 |
| Superior temporal | -0.027 | 0.017 | 0.19 | -0.06 | 0.006 | -0.026 | 0.017 | 0.22 | -0.06 | 0.008 | -0.065 | 0.019 | **0.0026** | -0.102 | -0.028 |
| Transverse temporal | 0.009 | 0.009 | 0.44 | -0.009 | 0.027 | 0.013 | 0.009 | 0.29 | -0.006 | 0.031 | 0.007 | 0.01 | 0.64 | -0.014 | 0.027 |
| Insula | 0.039 | 0.027 | 0.25 | -0.041 | 0.092 | 0.077 | 0.028 | **0.018** | 0.022 | 0.13 | 0.116 | 0.031 | **<0.0001** | 0.056 | 0.18 |
| Thalamus | 0.01 | 0.023 | 0.76 | -0.036 | 0.056 | 0.009 | 0.024 | 0.8 | -0.039 | 0.056 | 0.06 | 0.027 | **0.057** | 0.007 | 0.11 |
| Caudate | -0.061 | 0.017 | **0.002** | -0.095 | -0.027 | -0.095 | 0.018 | **<0.0001** | -0.13 | -0.06 | -0.097 | 0.02 | **<0.0001** | -0.14 | -0.058 |
| Putamen | -0.089 | 0.021 | **0.001** | -0.13 | -0.048 | -0.089 | 0.022 | **0.00032** | -0.13 | -0.046 | -0.092 | 0.024 | **0.0008** | -0.14 | -0.044 |
| Pallidum | -0.004 | 0.029 | 0.94 | -0.06 | 0.052 | -0.039 | 0.03 | 0.29 | -0.098 | 0.019 | 0.003 | 0.033 | 0.95 | -0.061 | 0.067 |
| Accumbens | -0.036 | 0.029 | 0.32 | -0.093 | 0.021 | -0.066 | 0.03 | **0.06** | -0.13 | -0.007 | -0.104 | 0.033 | **0.006** | -0.169 | -0.039 |
| Hippocampus | -0.038 | 0.023 | 0.18 | -0.084 | 0.008 | -0.004 | 0.024 | 0.93 | -0.052 | 0.043 | -0.032 | 0.027 | 0.34 | -0.084 | 0.02 |
| Amygdala | -0.006 | 0.034 | 0.93 | -0.072 | 0.06 | -0.057 | 0.035 | 0.19 | -0.13 | 0.012 | -0.161 | 0.038 | **0.0002** | -0.24 | -0.086 |
| Substantia Nigra | 0.005 | 0.35 | 0.94 | -0.064 | 0.073 | 0.045 | 0.036 | 0.33 | -0.026 | 0.12 | -0.024 | 0.04 | 0.66 | -0.101 | 0.054 |

**Supplementary Table 2. Progression of regional brain atrophy in PD.** Longitudinal atrophy changes measured using linear mixed effect models in regard to baseline. The significance threshold reported here is presented after FDR correction for multiple comparisons. Coeff = coefficient; FDR = false discovery rate; SE = standard error; CI = 95 % confidence interval.

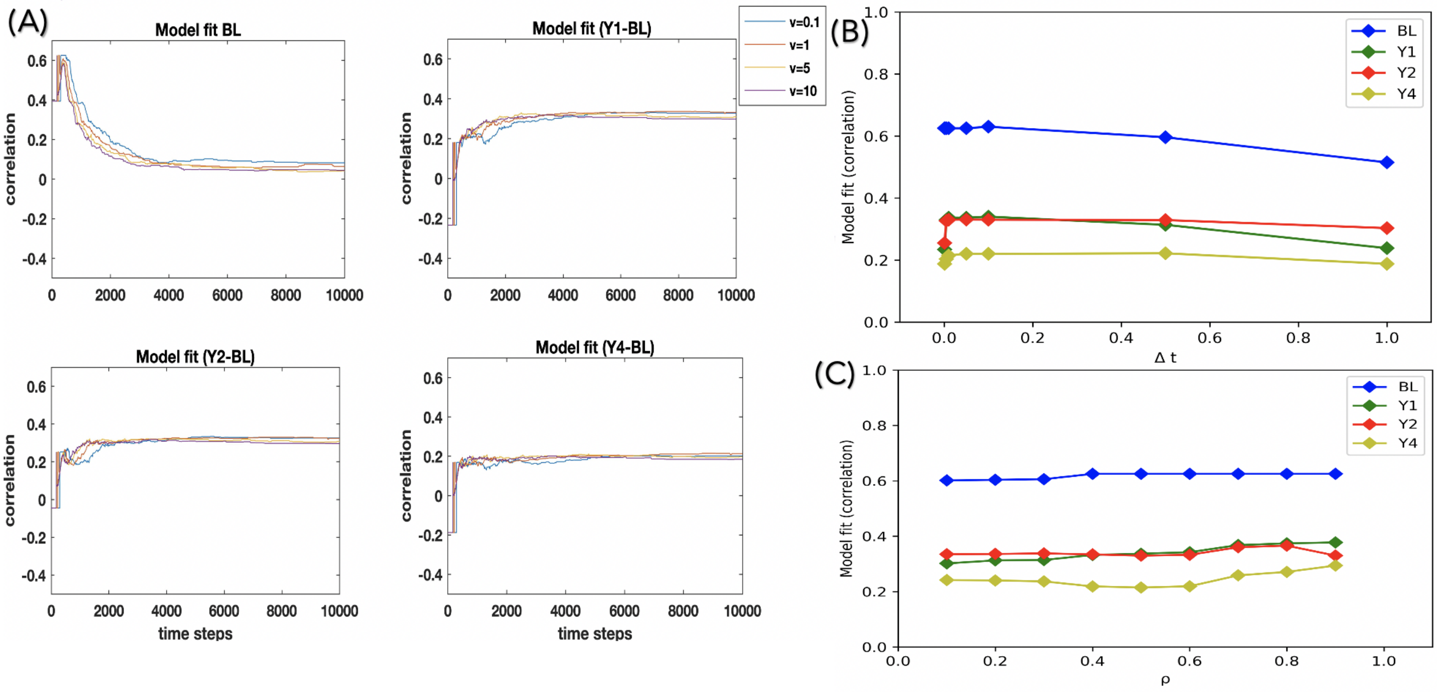


**Supplementary Figure 1. Effect of different model parameters.** The model fit measured using Spearman's rank correlation coefficients is robust to variations in **(A)** the propagation speed (v), tested using values ranging from 0.1 to 10 (v=1 in the main text), **(B)** the timestep increment ($\Delta t$), tested using values ranging from 0.001 to 1 ($\Delta t = 0.01$ in the main text), and **(C)** the probability of an agent staying in region *i* ($\rho$), tested using values ranging from 0.1 to 0.9 ($\rho$ = 0.5 in the main text) at the connection density of 35% used for main results. All parameters were tested at each time point, with distinct lines indicating peak correlation fits at baseline, at baseline versus one year, at baseline versus two years, and at baseline versus four years of follow-up.
